# Racial segregation, testing sites access, and COVID-19 incidence rate in Massachusetts, USA

**DOI:** 10.1101/2020.07.05.20146787

**Authors:** Tao Hu, Han Yue, Changzhen Wang, Bing She, Xinyue Ye, Regina Liu, Xinyan Zhu, Shuming Bao

## Abstract

The U.S. has merely 4% of the world population but 25% of the world’s COVID-19 cases. Massachusetts has been in the leading position of total cases since the outbreak in the U.S. Racial residential segregation is a fundamental cause of racial disparities in health. Moreover, disparities of access to health care have a large impact on COVID-19 cases. Thus, this study estimates racial segregation and disparities in testing sites access and employs economic, demographic, and transportation variables at the city/town level in Massachusetts. Spatial regression models are applied to evaluate the relationships between COVID-19 incidence rate and related variables. This is the first study to apply spatial analysis methods across neighborhoods in the U.S. to examine the COVID-19 incidence rate. The findings are: 1) residential segregations of Hispanic and Non-Hispanic Black/African Americans have a significantly positive association with COVID-19 incidence rate, indicating the higher susceptibility of COIVD-19 infections among minority; 2) The Black has the shortest drive time to testing sites, followed by Hispanic, Asian, and Whites. The drive time to testing sites is significantly negatively associated with the COVID-19 incidence rate, implying the importance of testing location being accessed by all populations; 3) Poverty rate and road density are significant explanatory variables. Importantly, overcrowding represented by more than one person per room is a significant variable found to be positively associated with COVID-19 incidence rate, suggesting the effectiveness of social distancing for reducing infection; 4) Different from previous studies, elderly population rate is not statistically significant with incidence rate because the elderly population in Massachusetts is less distributed in the hot spot regions of COVID-19 infections. The findings in this study provide useful insights for policymakers to propose new strategies to contain the COVID-19 transmissions in Massachusetts.

## 1. Introduction

The COVID-19 pandemic has severely impacted the socioeconomic activities worldwide since its outbreak in January. As of July 3, there have been 10,719,946 confirmed cases globally, including 517,337 deaths. The United States is the leading country with 2,671,220 confirmed cases and 127,858 deaths (WHO, 2020). Since the beginning of April, the US has become the COVID-19 pandemic center and the number of cases is still increasing. Social distancing is one of the most effective ways to reduce COVID-19 infection, but due to residential segregation, the separation of people based on income and/or race, some people from ethnic minority groups cannot practice social distancing. Hence, they are often found in overcrowded urban housing areas and make physical distancing and self-isolation difficult, thus leading to the increased risk for the spread of COVID-19 (Bhala, 2020). In addition, socioeconomic inequities frequently impact health and healthcare access, resulting in a higher burden of disease and mortality in vulnerable social groups (Daniel, 2020). Therefore, it is necessary to integrate social-economic information and disease statistics to help analyze and understand the spread of COVID-19.

Many research findings have highlighted the racial disparities in COVID-19 infections. Across the country, deaths caused by COVID-19 are disproportionately high among African Americans (Dorn, 2020), while Chicago and New York City reported greater COVID-19 mortality among Latinos (Hooper, 2020). To utilize more detailed information on the racial and socioeconomic disparities, Matthew and Julia (2020) used 2018 Behavioral Risk Factor Surveillance System (BRFSS) to estimate the proportion of adults that have at least one CDC criteria for risk of severe illness from COVID-19. The analysis is categorized by age group, race, and household income. Results show that people who are Black, American Indian, or live in low-income households are more likely to have conditions associated with increased risk of illness from COVID-19 compared to those who are white or have a higher income, respectively. Anyane□Yeboa, et al. (2020) investigated the racial disparity of infection and deaths caused by COVID-19 in the US and assessed the rates of COVID-19 infection and death by race and ethnicity with information obtained from the Department of Health websites of sixteen states. They stated Black patients had higher rates of infection and death from COVID-19, which are consistent with the findings from some other individual states. Cato and Aneesah (2020) presented the overview of racial and ethnic distribution of COVID-19 confirmed cases and fatalities in the state of Connecticut to demonstrate the unique challenges among Black and Brown communities. At the metropolitan level, Yu et al. (2020) examined the growth rate of both COVID-19 confirmed cases and deaths in the first 30-day period of the outbreak in the 100 largest metropolitan cities and observed that the growth curve was particularly steep in counties that are located in cities with high economic disparity and residential segregation of Blacks and Hispanics.

One fundamental cause of racial disparities in health is racial residential segregation, which presents the physical isolation of one racial group from others (Williams and Collins, 2001). This segregation can affect health through concentrated poverty, the quality of the neighborhood environment, and the individual socioeconomic attainment of minorities (Dolores and Locher, 2003). Health experts believe that person-to-person and community transmission are the most common ways to spread the COVID-19. Thus, neighborhoods with concentrated poverty and over-occupancy of housing units are at a higher risk of COVID-19 infection. Moreover, minorities under the poverty line are more likely to work in industries that have remained open during non-essential business closures (Raifman, 2020). Thus, they have greater exposure to COVID-19 and contribute to the transmission of COVID-19 as well. Given the influence of racial residential segregation on the socioeconomics of neighborhoods, it is necessary to conduct a systematic investigation of whether segregation has a direct impact on the spreading of COVID-19.

In addition, adequate access to affordable testing sites and hospitals is critically important to identify potential carriers of COVID-19. Since it is the best way to provide evidence-based decisions to slow down the disease, the World Health Organization (2020) has called on countries to ‘test, test, test’ coronavirus. Thus, appropriate measures, such as isolation and hospitalization, can be taken to contain the pandemic. Many states, with the support of the federal government, have expanded access to COVID-19 related health care services either by setting up more mobile testing sites and increasing testing providers or providing telehealth to cover as many people as possible. However, these testing sites are more likely to be distributed in the well-off suburbs of white-dominant neighborhoods rather than in low-income minority neighborhoods (Williams and Cooper, 2020). Such disparities in health care access are becoming worse among socioeconomically disadvantaged groups due to the lack of health insurance, access to transportation, and individual awareness of the disease severity. There has been an estimated 25% of the black population accounting for 41% of COVID-19 cases in Boston (Jonas, 2020). Also, the African American and Hispanic communities are reported to be at higher risk of infection and death from COVID-19. Hence, a reliable and accurate measure of access to testing sites could help us understand which areas and what demographic groups suffer inadequate access and what testing strategies may be adopted to mitigate the COVID-19 pandemic.

According to previous studies, socio-demographic and economic, and environmental features are also important factors in affecting the spread of COVID-19 disease. For example, in the analysis conducted by Mollalo et al. (2020), they considered income inequality, median household income, the percentage of nurse practitioners, and the percentage of the black female population when modeling the COVID-19 incidence at the US county level. The results demonstrated that areas with a high incidence of COVID-19 usually have high-income inequality and median household income (Mollalo, 2020). Liu (2020) collected the number of laboratory-confirmed COVID-19 cases in 312 cities in China, and a series of sociodemographic variables such as distance to the epicenter, the total length of built urban metro lines, urban area, population density, the annual quantity of wastewater discharged, and residential garbage connected and transported, per capita public recreational green space, the daily highest temperature, and the capital city. Based on these data, a study of the impacts of COVID-19 transmission was conducted from the urban perspective. The statistically significant results revealed that residential garbage connected and transported, and the annual quantity of wastewater discharged could increase the confirmed infection number of COVID-19 (Liu, 2020).

To explore regional patterns and inform local policymakers about the efficient allocation of resources and personnel to e□ectively mitigate the spread of Covid-19, making inferences at finer scales, such as the sub-county, census tract, or block group, may produce more accurate results than coarse levels, such as the county or above. In Chicago, more than 50% of COVID-19 cases and nearly 70% of COVID-19 deaths involve black individuals. These deaths are concentrated in just 5 neighborhoods in the city’s South Side. Thus, it is critical to quantify the interaction between the various factors and disease statistics at a finer scale. Analyzing regional patterns and increasing their awareness of the COVID-19 dangers and preventative measures, will benefit the most a□ected communities (Daniel, 2020). Quantifying disparities in risk is important for allocating resources to prevent, identify, and treat COVID-19-related severe illness and limit diverging outcomes for vulnerable subgroups.

The research aims to identify the impact of racial segregations and testing sites accessibility on the COVID-19 incidence rate in cities/towns of Massachusetts. To the authors’ knowledge, it is the first study to employ spatial analysis methods on neighborhoods’ COVID-19 data in the US. The objectives are: (1) evaluating the minority racial segregation, such as Hispanics, Non-Hispanic Black Americans, and Non-Hispanic Asians and its socioeconomic characteristics in Massachusetts; (2) accessing the spatial accessibilities to testing sites across different sociodemographic groups in the study area; (3) investigating whether neighborhoods with higher COVID-19 incidence rate are positively associated with highly segregated areas for minority ethnics; (4) exploring whether testing sites is well distributed for the COVID-19 testing; and (5) examining the association between socioeconomic and COVID-19 incidence rate.

## 2. Study area and data

### 2.1 Study Area

Massachusetts has an estimated population of 6,547,785 million as of 2019 according to the US Census Bureau. Currently, Massachusetts is the fifteenth-most populous U.S. state with small fluctuation in recent years. For the racial constitutions in Massachusetts, 80% of the population are White alone, 9% are Black or African American alone, 7.2% are Asia alone, and other racial populations are 3.9%. 63.4% of the population are in the age group of 18 and 64, while 17% are in the age group of 65 and over (US Census Bureau, 2020). There are 357 cities/towns in Massachusetts and 4 of them are undefined areas according to the base map provided by the US Census Bureau. Massachusetts is composed of Eastern, Central, and Western regions. Eastern Massachusetts is more urban than Western Massachusetts. Boston is the largest city, at the inmost point of Massachusetts Bay. Worcester County is usually considered to be in central Massachusetts. The center of the population of Massachusetts is in Middlesex County, in the town of Natick.

As of May 20, Massachusetts has 88,970 confirmed cases leading in the 5th place in the US and 6,066 deaths ranking as the 3rd place. For the total confirmed cases, 29.6% are White, 19.1% are Hispanic, and 9.4% are Black/African American. In terms of the reported hospitalization, the percentage of Non-Hispanic White is up to 48.1%, Hispanic and Black/African Americans are 13.3% and 11.6% respectively. These numbers suggest the White population being infected with COVID-19, has a higher rate of hospitalization than that of Hispanic and Black/African American. Thus, analysis of the racial and health care access disparities is needed in Massachusetts.

On March 10, Massachusetts declared a state of emergency, taking steps to limit the spread of COVID-19. The Department of Public Health (2020a) provides not only daily and cumulative reports on Massachusetts COVID-19 cases, testing, and hospitalizations but also weekly and biweekly reports including nursing facility data, cases by city/town, residents subject to COVID-19 quarantine, and data from State facilities. The city/town COVID-19 incidence rate (cases per 100,000 people) data is recorded from April 14. Massachusetts government (2020a) also publishes testing site locations in the state and we geocode the location to latitude and longitude. Figure 1 demonstrates the spatial distribution of the COVID-19 incidence rate and testing sites as of May 20, 2020. There is an extremely uneven distribution pattern: cities/towns with high incidence rates were aggregated in the Greater Boston area. In the central and west of Massachusetts, a few places are the hotspot of high incidence rates.

**Figure1.**
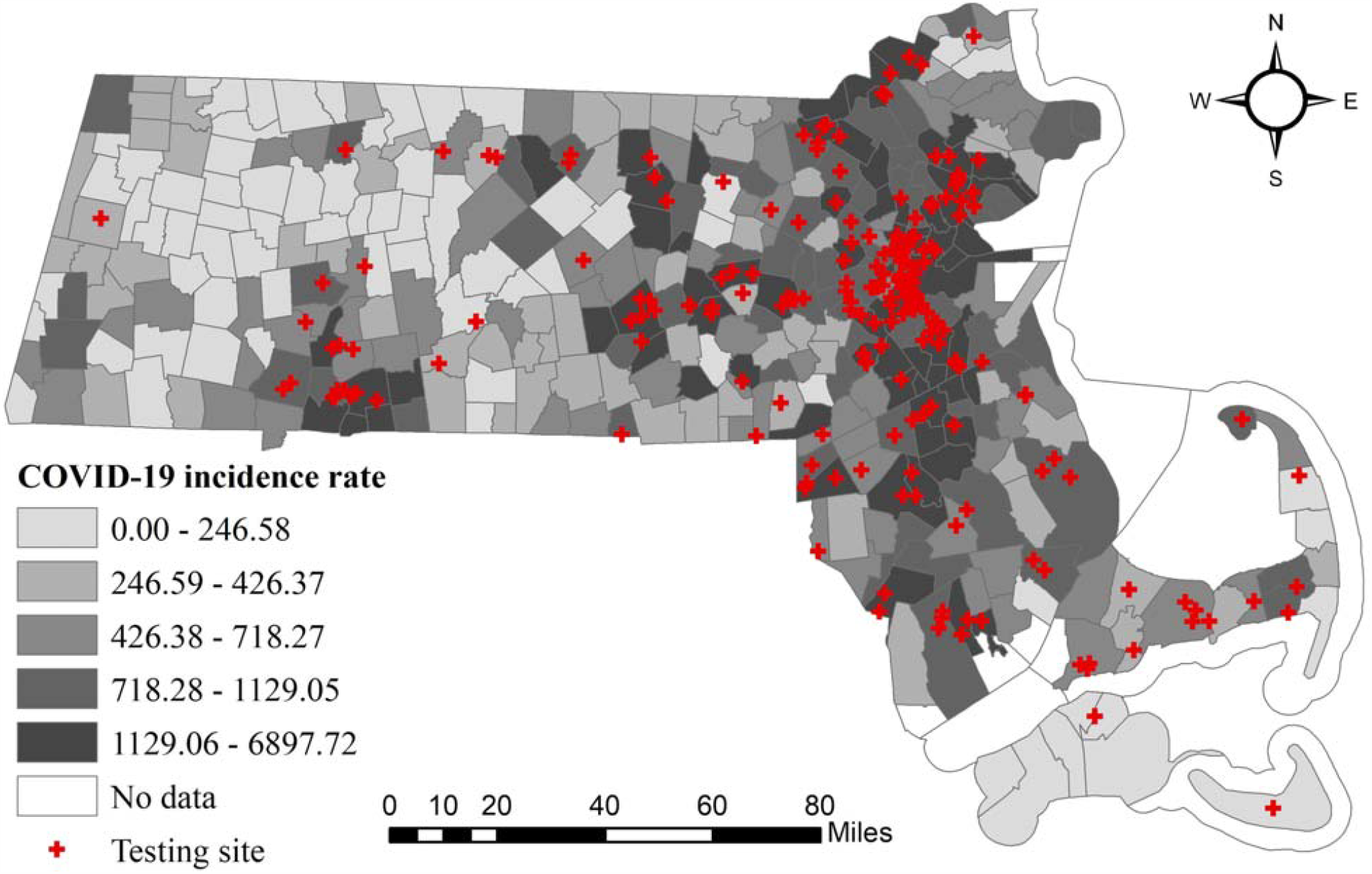
City/town-level accumulated COVID-19 incidence rate (confirmed cases/per 100,000 people) and testing sites in Massachusetts as of May 20, 2020.

### 2.2 Data Preparation

This study aims to investigate the relations between COVID-19 incidence rate and racial residential segregation, access to testing sites, and other socio-demographic and economic variables. These data are gathered from different sources. The city/town COVID-19 incidence rate data are collected from the Department of Public Health in Massachusetts. The racial residential segregation and disparities in testing site accessibilities are compiled from racial population data in cities/towns and sub-unit, census block groups. There have been many studies involving various socioeconomic and demographic factors that may impact the COVID-19 incidence rate, including the elderly population rate, poverty rate, overcrowding rate, household income, and so on. We collect such variables both from the subcounty and census block group (CBG) 2018 American Community Survey (ACS) provided by the US Census Bureau (https://www2.census.gov/geo/tiger/TIGER_DP/2018ACS/).

This study estimates minorities’ residential segregations, including Hispanic, Non-Hispanic Black or African Americans, and Non-Hispanic Asian. To simplify the name of each race, we use Black and Asian to present Non-Hispanic Black or African American and Non-Hispanic Asian in the rest of the paper. Residential segregation can be discussed in five distinct dimensions: centralization, concentration, clustering, unevenness, and isolation (Massey and Denton, 1988). This study applies the isolation index to associate segregation and health outcomes. The isolation index can effectively reveal the racial differential sizes (Chang, 2006).

Also, it is different from the racial percentage, which is hard to tell the racial distribution disparities. The higher of the racial isolation index indicates the same racial population has a higher chance to live as neighbors. It is a vital variable to study COVID-19 transmission patterns. Thus, this research applies the isolation index to estimate the segregation of each race. The isolation index works as follows. Assuming city/town *j* consists of *n* census block groups, the isolation index for a race within j can be presented as:

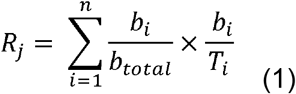

where *i* is the *i*-th census block group in the city/town j;*b*_*i*_ is the race population in i;*b*_*total*_ is the total race population in *j*, and *T*_*i*_ is the total population in *i*. The isolation index ranges from 0 to 1.0 indicates no segregation and 1 indicates the greatest segregation. The index presents the chance of having the same race live as neighbors.

Access to testing sites is measured by proximity, a popular method in geographic information systems (GIS), and the most influential component in health-related studies. That is, the shortest travel times to the nearest testing sites and hospitals respectively. Both the testing site providers and primary care physicians in the hospitals are the first contact with probable COVID-19 patients. One issue is people may not go to the closest site for testing whether they are infected COVID-19 due to various reasons, such as long waiting time and no available providers. They may divert to the secondary choice of traveling a longer distance. Another issue might be the omission of spatial accessibility accounting for both supply and demand. Due to the unknown capacity of testing sites and hospitals for COVID-19, such two travel times are still acceptable to estimate access to COVID-19 related health care services. We use the population-weighted centroids of 357 cities/towns calibrated from the 2010 census block group level data to represent the location of the population in Massachusetts. Compared to the geographic centroids, the population-weighted centroids have better accuracy, particularly in rural or peripheral suburban areas (Wang, 2015: 78), thus are commonly used. The location information of testing sites is first downloaded from the Massachusetts Government website (2020b) and then geocoded by Google Maps. We also geocode the location of hospitals extracted from COVID-19 Weekly Public Health Report (Massachusetts Government Website, 2020a). There are 228 testing sites and 76 hospitals in Massachusetts. Note that there might be some people crossing the state’s border to neighboring states or some other people outside of the state coming for testing. We don’t consider these two cases. The travel times to testing sites and hospitals are estimated through Google Maps Distance Matrix API as it represents the dynamic traffic conditions and routing rules (Wang and Xu, 2011), which are not captured by the simplified Euclidean or road network-based distance accounting for the speed limits. In addition, we calculate the weighted average travel times based on the population across four demographic groups.

Overall, the data used in this study are in four categories: racial segregation, accessibility, demographics, economics, and transportation. In terms of demographic, elderly population rate (percent of 65 years and over), overcrowding rate (percent of more than 1 occupants per room), and education attainment are estimated. The economic variables include poverty rate, household income, and income inequality. Transportation includes road density and public transit rate (percentage of workers 16 years and over who go to work by public transportation (excluding taxicab)). Table 1 presents the name, description, and source of the estimated index and variables described above.

**Table 1.** Definition, sources, and summary statistics of explanatory variables used in this study.

| Category | Variable Name | Definition | Source | Mean | Std. Dev. | Min | Max |
| --- | --- | --- | --- | --- | --- | --- | --- |
| Race segregation | (1) Segregation of Hispanic | Isolation index of Hispanic | American Community Survey 2018 | 0.87 | 0.12 | 0 | 1 |
|  | (2) Segregation of Non-Hispanic Black/African American | Isolation index of Black/African American |  | 0.06 | 0.07 | 0 | 0.53 |
|  | (3) Segregation of Non-Hispanic Asian | Isolation index of Asian |  | 0.06 | 0.07 | 0 | 0.42 |
| Accessibility | (4) Travel time to testing sites | The drive time from the population-weighted centroid of each city/town to the nearest testing sites (minute) | Google Maps Distance Matrix API | 12.67 | 9.27 | 1 | 57 |
|  | (5) Travel time to hospitals | The drive time from the population-weighted centroid of each city/town to the nearest hospitals (minute) |  | 15.68 | 8.60 | 1 | 61 |
| Demographic | (6) Percent of 65 years and over | Percentage of the population over 65 years old (%) | American Community Survey 2018 | 18.22 | 5.83 | 7.50 | 43.07 |
|  | (7) Percent of more than 1 occupants per room | Percentage of occupied housing units with more than 1 occupants per room (%) |  | 1.18 | 1.24 | 0 | 9.28 |
|  | (8) Education attainment | Percentage of population (25 years and over) with education level no less than bachelor's degree (%) |  | 42.94 | 16.12 | 11.31 | 84.73 |
| Economic | (9) Poverty rate | Percentage of population with income in the past 12 months below poverty level (%) |  | 7.37 | 4.76 | 0.54 | 33.23 |
|  | (10) Household income | Median household income in the past 12 months (in 2017 inflation-adjusted dollars) (dollar) |  | 85700.6 | 28662.39 | 0 | 204018 |
|  | (11) Income inequality | Gini index of household income |  | 0.43 | 0.05 | 0.31 | 0.57 |
| Transportation | (12) Public transit | Percentage of workers 16 years and over who go to work by public transportation (excluding taxicab) (%) |  | 4.13 | 5.96 | 0 | 33.64 |
|  | (13) Road density | The total length of primary and secondary roads for each city/town calculated/area of the corresponding city/town (per km) | US Census Bureau TIGER/Line | 2.87 | 2.31 | 0 | 13.73 |

## 3. Methodology

In this study, we first use Moran’s I index to test the spatial autocorrelation pattern of the city/town-level COVID-19 incidence rate in Massachusetts. Then, two classical spatial autoregressive models, including the spatial lag model (SLM) and the spatial error model (SEM), are adopted to determine the factors that influence the COVID-19 incidence rate.

### 3.1 Spatial autocorrelation test

Compared with geographical events that are far apart from each other, neighboring geographical events tend to share similar conditions, which makes them more closely related to each other than distant events (Tobler, 1970). This phenomenon is referred to as spatial autocorrelation or spatial dependency, which indicates whether the distribution of data depends on geographical locations (Cliff & Ord, 1973). If data at adjacent locations are similar to each other, the spatial autocorrelation effect is regarded as positive. If data at adjacent locations differ greatly from each other, the spatial autocorrelation effect is negative. If the distribution of data is irrelevant to geographical locations, there is no obvious spatial autocorrelation effect. A commonly used indicator to identify the spatial autocorrelation effect is Moran’s I index (Moran, 1948), which is formulated as:

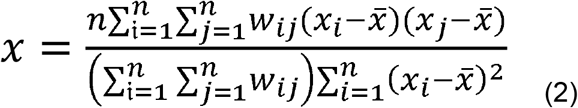

where *x*_*i*_and *x*_*j*_ mean the attribute of *i-*th and *j-*th spatial unit (the COVID-19 incidence rate of a city/town in this study), 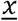 indicates the average of all the attributes, *n* means the number of spatial units, and *w*_*ij*_ is a member of the spatial weight matrix *W* which represents the spatial relationship between spatial unit *i* and *j*. In this research, we adopted the frequently-used rook method to construct the weight matrix, i.e., if spatial unit *i* and *j* share a common border,then *w*_*ij*_ =1, otherwise, *w*_*ij*_ =0 (Anselin, 1988).

The value of Moran’s I index ranges between -1 and 1: value less than 0 means a negative spatial autocorrelation effect, and the smaller the value is, the stronger the negative spatial autocorrelation effect is; a value greater than 0 indicates a positive spatial autocorrelation effect, and the larger the value is, the stronger the positive spatial autocorrelation effect is; and a value equals to 0 means no spatial autocorrelation effect. In addition to this, the Z-value can be applied to assess the statistical significance of the Moran’s I index, i.e., if the Z-value is larger than 1.96 or smaller than -1.96, then there is a spatial autocorrelation effect with a confidence level of 95%.

### 3.2 Spatial regression model

Spatial data usually presents a certain degree of positive spatial autocorrelation, i.e., attributes of adjacent geographical events tend to be more similar than attributes of geographical events that are further away from each other (Anselin & Bera, 1998). The existence of spatial autocorrelation effect violates the basic independent identical distribution (IID) assumption of classical regression models, i.e., observations are independent of one another. The significance of estimates will be overestimated if this effect is not taken into account (Clifford & Richardson, 1985). In this study, we applied two common spatial regression models to tackle the spatial autocorrelation effect, including the spatial lag model (SLM) and the spatial error model (SEM). The SLM and SEM account for the spatial autocorrelation effect in different ways: in the SLM, the dependent variable at a location is influenced by dependent variables of neighboring locations. While in the SEM, the spatial autocorrelation effect is represented through the remainder terms, i.e., the error at a location is affected by errors from neighboring locations.

The SLM can be formulated in terms of COVID-19 incidence rate:

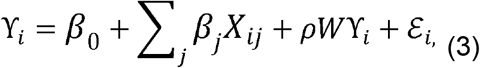

where *i* denotes a city/town, γ _*i*_ indicates the dependent variable (COVID-19 incidence rate) of the *i*-th city/town, *x*_*ij*_ means the *j*-th explanatory variable of city/town *i*, β* are unknown parameters to be estimated which measure the association between COVID-19 incidence rate and covariates *ceteris paribus, w*γ_*i*_ is the spatial lag variable in which *W* is a spatial weight matrix, ρ is the spatial autoregressive coefficient that indicates the spatial dependence of the explained variable, and *ε*_*i*_ is a random error term.

The SEM is formulated as:

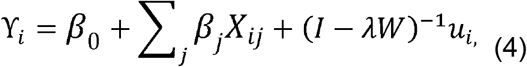

where *I* is an identity matrix, λ is the spatial autoregressive coefficient that represents the spatial dependence of residuals, and *u*_*i*_, is a normally distributed error term. The remaining symbols have the same meaning as those in Formula 2.

As shown in Table1, the covariates have different units of measurement and large disparities in magnitude, we standardized these variables to have a mean of 0 and a variance of 1. This could make the parameter estimates independent of units and easy to compare.

## 4. Results

### 4.1 Exploratory Analysis

The COVID-19 incidence rate varies across different socio-demographic groups at the city/town level. As shown in Table 2, the Black population has the lowest mean value of 2.75% ranging from 0 to 43.95%, followed by Asian and Hispanic populations with mean values of 3.6% and 4.83%. The white population has the highest mean value of 91.06% ranging from 39.27% to 100%, reflecting the dominance of white people in Massachusetts. These four groups experience very similar COVID-19 incidence rates across the first three quantiles and then the rate differences enlarge in the fourth quantile. Using the first quantile as a reference, Figure 2 shows the variation of the COVID-19 incidence rate ratio of each quantile with that of the first quantile. All four groups exhibit a smooth increase with minor differences and then the ratio dramatically moves up to a range of 9 to 16. Due to the large amount of confirmed COVID-19 cases in Boston city, we recalculate the COVID-19 incidence rate in the fourth quantile outside of Boston city in Table 2 and Figure 3. The rate ratios of Asian and White are consistent with those in the entire study area while Black and Hispanic groups exhibit a small increase in incidence rate, which are aligned with the concentrated segregation indices of Black and Hispanic in Boston city in Figure 2.

**Table 2.**
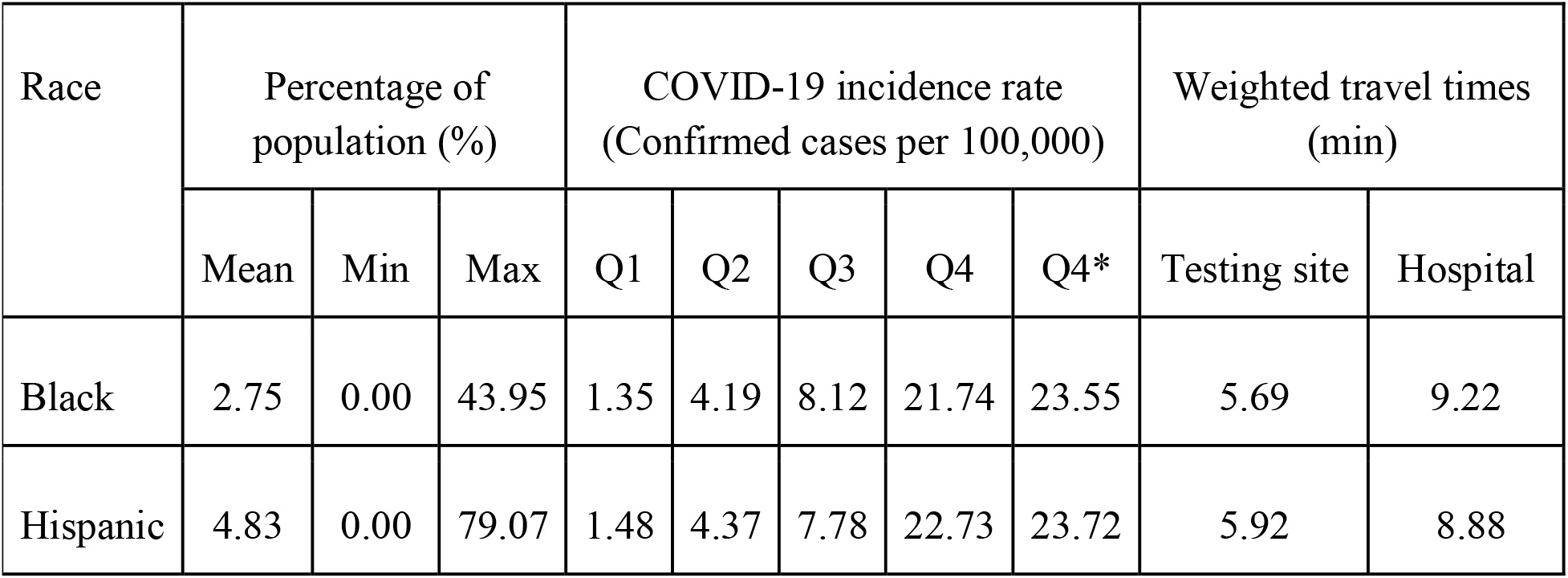
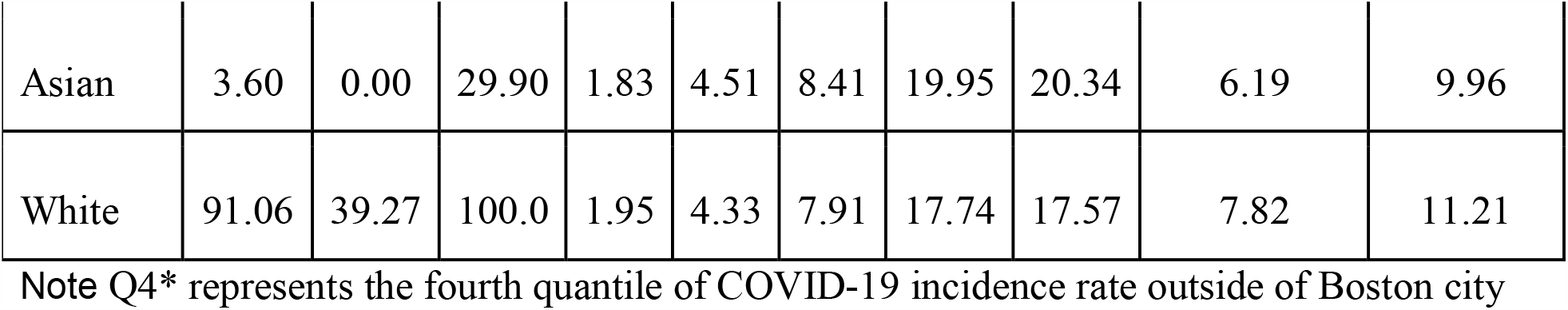
COVID-19 incidence rate and health access across racial population in city/town level.

| Race | Percentage of population (%) |  |  | COVID-19 incidence rate<br>(Confirmed cases per 100,000) |  |  |  |  | Weighted travel times<br>(min) |  |
| --- | --- | --- | --- | --- | --- | --- | --- | --- | --- | --- |
|  | Mean | Min | Max | Q1 | Q2 | Q3 | Q4 | Q4* | Testing site | Hospital |
| Black | 2.75 | 0.00 | 43.95 | 1.35 | 4.19 | 8.12 | 21.74 | 23.55 | 5.69 | 9.22 |
| Hispanic | 4.83 | 0.00 | 79.07 | 1.48 | 4.37 | 7.78 | 22.73 | 23.72 | 5.92 | 8.88 |
| Asian | 3.60 | 0.00 | 29.90 | 1.83 | 4.51 | 8.41 | 19.95 | 20.34 | 6.19 | 9.96 |
| White | 91.06 | 39.27 | 100.0 | 1.95 | 4.33 | 7.91 | 17.74 | 17.57 | 7.82 | 11.21 |

Note Q4\* represents the fourth quantile of COVID-19 incidence rate outside of Boston city

**Figure 2.**
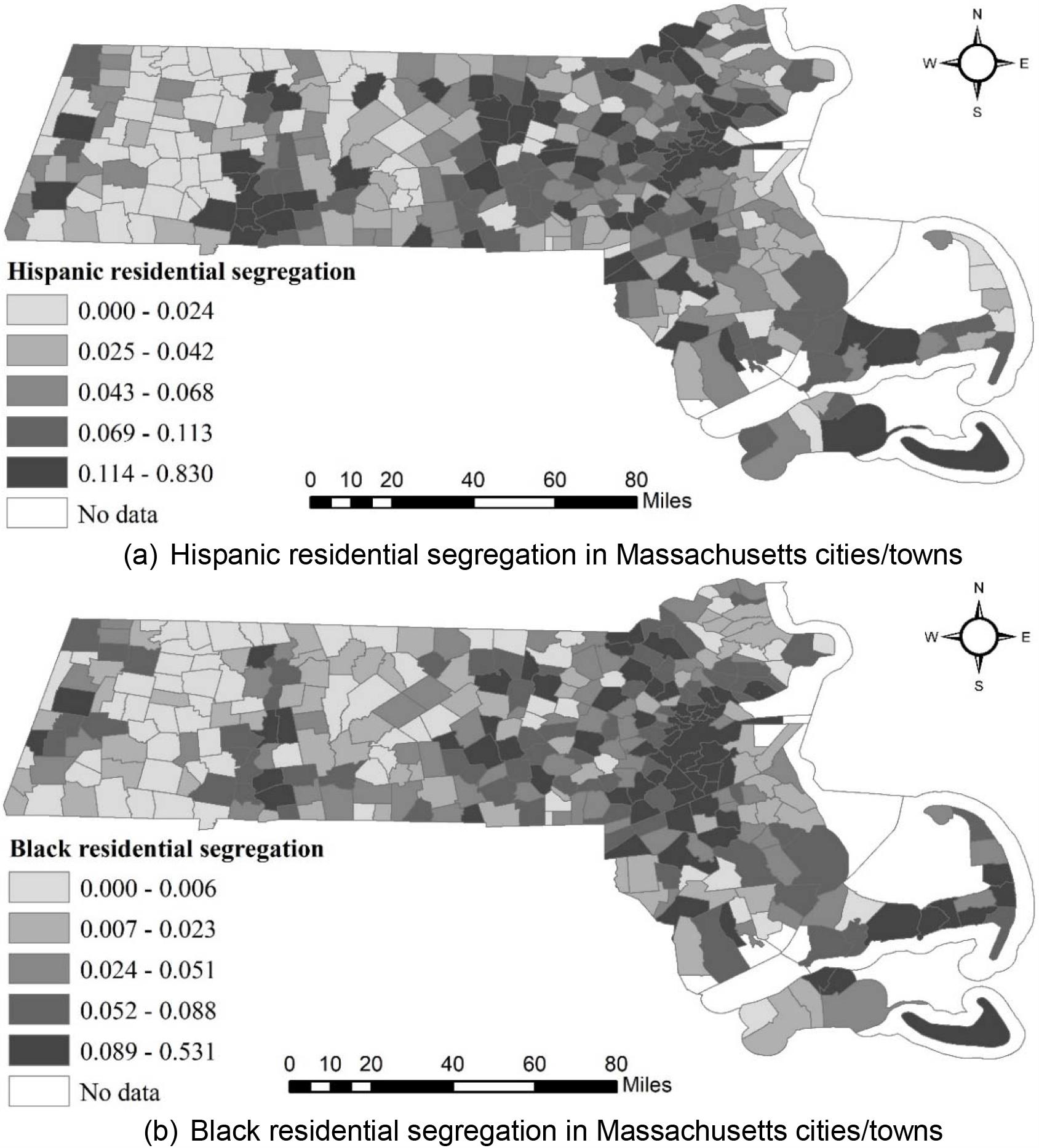
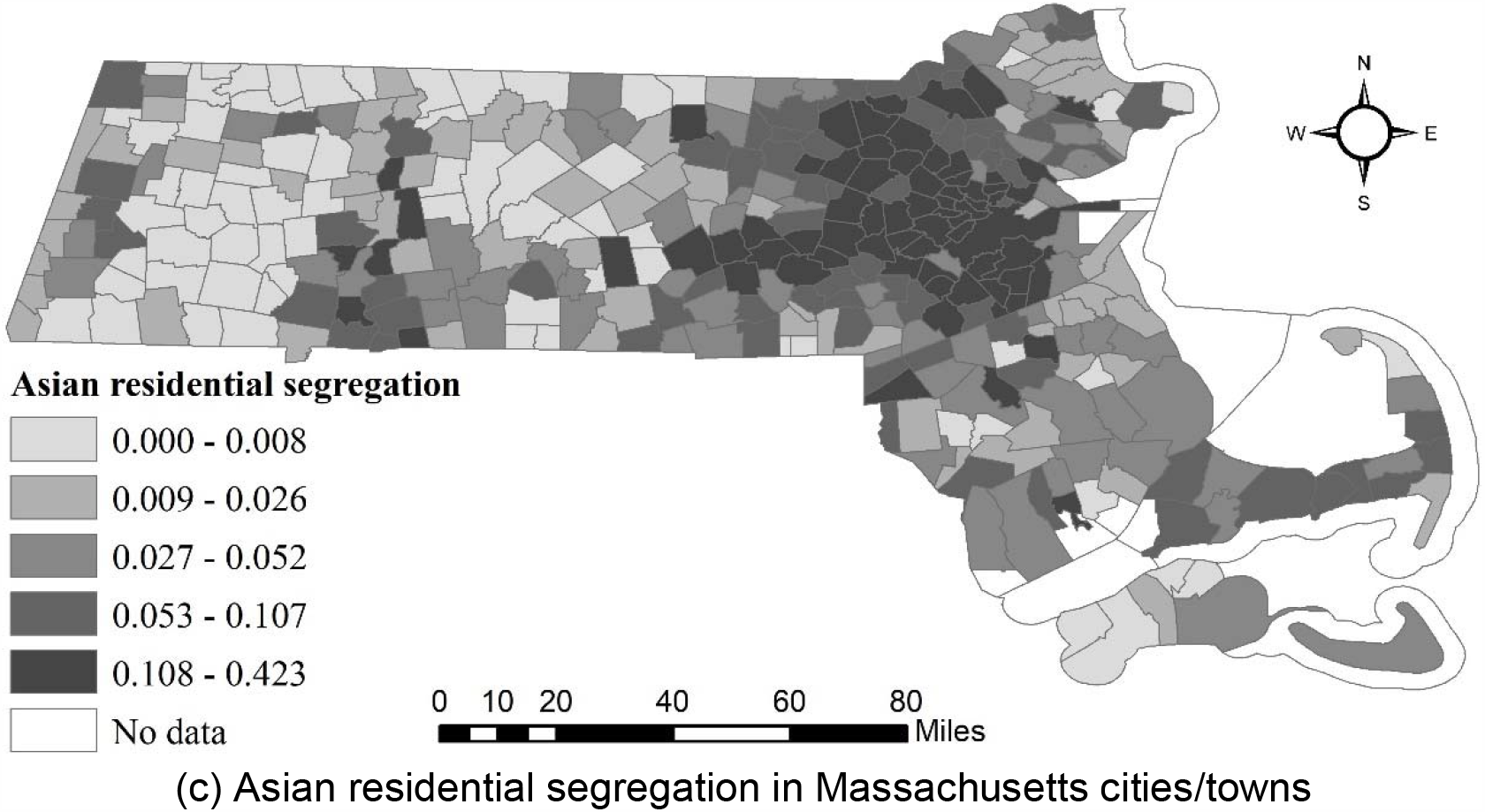
Racial segregation index of Hispanic, Non-Hispanic Black, and Non-Hispanic Asian.

**Figure 3.**
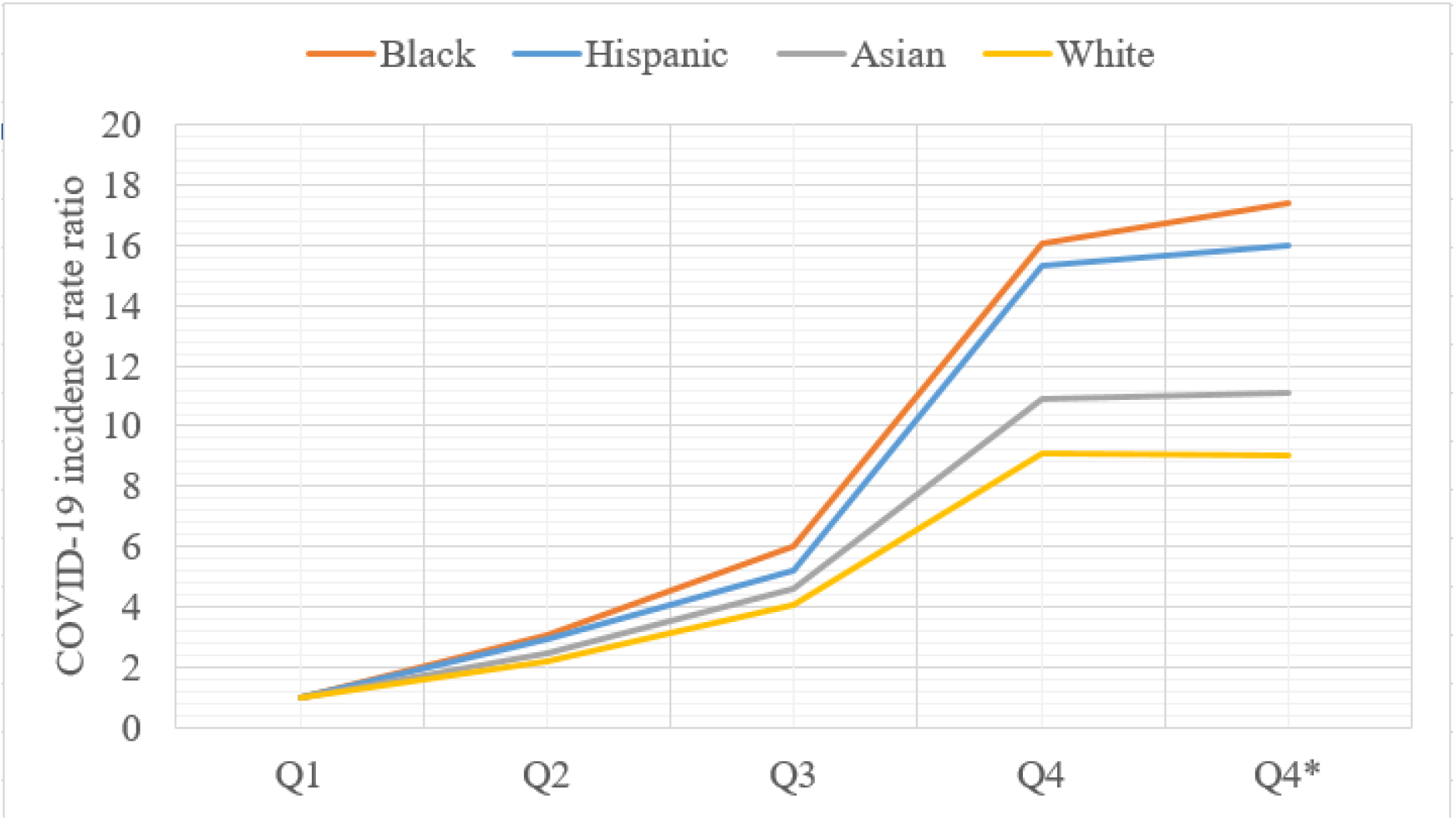
Variation of COVID-19 incidence rate across racial population in city/town, MA. The COVID-19 incidence rate ratio represents the ratio of the COVID-19 incidence rate in each quantile compared to the first quantile.

Racial residential segregation is estimated by the isolation index described in Section2. Figure 2 illustrates the segregations of the minorities in Massachusetts, including Hispanic, Black, and Asian. Higher values indicate higher possibilities that people of the same race are living as neighbors. As shown in Figure 2, Hispanic and Black in Boston have higher segregation indexes than Asian. Meanwhile, Asians are more likely to live together at Middlesex and Norfolk. Middlesex County is amongst the top 25 counties with the highest household income and the 25 most populated counties.

To quantify the health disparities across different racial groups, the weighted travel times to testing sites and hospitals are illustrated in the last two columns of Table 2. The Black group has the lowest weighted travel time of 5.69 minutes to the testing sites, followed by the Hispanic, Asian, and White groups. Access to hospitals tells a similar story with a minor difference. The Hispanic group surpasses the Black with the lowest travel time of 8.88 minutes. Compared with the other three groups, the White group has the longest travel times of 7.82 and 11.21 minutes to testing sites and hospitals respectively, which is consistent with the lowest COVID-19 incidence rate in more concentrated quantiles. In addition, the travel times to testing sites and hospitals are both in acceptable ranges less than 15 minutes across each group. However, the travel times to testing sites is almost 3 minutes less than the travel times to hospitals, which revealed the condition of accessible health care services related to COVID-19 in Massachusetts.

### 4.2 Results of spatial regression analysis

We conducted a Moran’s I test to examine the spatial effects. The corresponding result implies that the spatial distribution of city/town-level COVID-19 incidence rate offers strong evidence of spatial autocorrelation with Moran’s I equals to 0.436, which is positive and significantly different from expected Moran’s I of -0.003 (*p*-value<0.001, z-score=13.196). The positive sign implies the existence of spatial adjacency effect. The COVID-19 incidence rate of a city/town affects that of other nearby cities/towns, partially because adjacent cities/towns share similar attributes. The *p*-value is considerably lower than 1%, indicating that the existence of spatial autocorrelation in the COVID-19 incidence rate is statistically significant.

After correlation analysis (correlation coefficients<0.6), among the 12 candidate explanatory variables, 9 variables were selected to be included in the final models. These variables are the percentage of elderly people (65 years and over), percent of more than 1 occupants per room, poverty rate, income inequality, road density, and test site accessibility. Table 3 shows the results of the two spatial regression models. For comparison, the classic OLS model was also calibrated, and results were listed.

**Table 3.** Results of regression models in town/city level.

| Variable | OLS | SLM | SEM |
| --- | --- | --- | --- |
| $\rho$ (spatial lag coefficient) | - | 0.433*** (0.050) | - |
| $\lambda$ (spatial error coefficient) | - | - | 0.521*** (0.059) |
| Constant | 728.148*** (23.496) | 413.507*** (42.601) | 718.326*** (43.502) |
| Segregation of Hispanic | 242.532*** (36.383) | 211.309*** (32.708) | 215.677*** (34.596) |
| Segregation of Non-Hispanic Black | 112.717*** (29.986) | 75.039*** (26.837) | 47.236* (27.690) |
| Segregation of Non-Hispanic Asian | -27.229 (32.422) | -37.502 (28.965) | -47.194 (31.288) |
| Test site accessibility | -64.349** (29.140) | -37.318 (26.405) | -75.207** (37.491) |
| Percent of 65 years and over | -0.359 (28.820) | 12.8947 (25.741) | 33.510 (32.452) |
| Percent of more than 1 occupants per room | 155.081*** (30.420) | 153.784*** (27.250) | 157.385*** (27.722) |
| Poverty rate | -93.610*** (32.056) | -39.326 (29.119) | -66.217** (31.659) |
| Income inequality | -29.444 (29.006) | -32.067 (25.936) | -17.368 (27.376) |
| Road density | 180.444*** (37.660) | 112.369*** (35.468) | 226.89*** (41.298) |
| Log-likelihood | -2621.59 | -2596.979 | -2594.24 |
| AIC | 5263.18 | 5213.96 | 5210.48 |
| $R^2$ | 0.579 | 0.654 | 0.657 |
| Moran's I of residuals | 0.436*** (z-score: 13.196) | 0.032 (z-score: 1.017) | 0.002 (z-score: 0.140) |

Note: \* Significant at 0.1; \*\* significant at 0.05 level; \*\*\* significant at 0.01 level. Standard errors are in parentheses. The diagnostic test demonstrated that the multicollinearity condition number is 4.587, which indicated that there was no serious multicollinearity.

Three metrics were used to compare the performances of the OLS, SLM, and SEM: log-likelihood at convergence (log-likelihood), Akaike information criterion (AIC), and *R*-squared (*R*^2^). For log-likelihood and *R*^2^, a higher value means better performance; for AIC, a lower value represents better performance. The results of the model performances suggested the following. First, two spatial models could better fit the observations than OLS could: the AIC of the OLS (5263.18) was much larger than in the SLM (5213.96) and SEM (5210.48), while the log-likelihood and *R*^2^ of the OLS (−2621.59, 0.579) were smaller than those of the SLM(−2596.979,0.654) and SEM (−2594.24,0.657). Second, the performance of the SEM was slightly better than that of the SLM: the log-likelihood and *R*^2^ of the SEM were larger than those of the SLM, while the AIC of the SEM was smaller than that of the SLM. The highest R^2^ is achieved by SEM (0.656657) that explains 65.67% of the total variations of COVID-19 incidence rates.

A tenet of regression is that residuals should be independent of each other, that is, they should be randomly distributed in space. Furthermore, the degree to which residuals are autocorrelated in space is another important indicator to judge the performance of regression models. The results showed that the residuals of the OLS revealed a strong positive autocorrelation, while the residual autocorrelations of the SLM and SEM were eliminated, and the residual autocorrelation of the SEM was eliminated more thoroughly than that of SLM (the z-score of SEM (0.140) is smaller than that of SLM (1.017).

According to the above-mentioned results, by incorporating spatial dependence, the two spatial regression models improve the performance of OLS in modeling the COVID-19 incidence rate in Massachusetts. Additionally, the SEM outperformed the SLM, which means that the spatial spillover effect in the data is mainly reflected in the residuals. Therefore, the SEM specification is a more appropriate choice in this research to account for the spatial autocorrelation effect.

The results of the three models demonstrate that segregations of Hispanic and Black are significantly positively associated with the COVID-19 incidence rate, while segregation of Asian had a negative and nonsignificant influence on the COVID-19 incidence rate. As to the results of SEM, a one-point increase in the segregation of Hispanic and segregation of Black is respectively associated with a 215.677-point, a 47.236-point increase in the COVID-19 incidence rate, while a one-point increase in the segregation of Asian is associated with a 47.194 decrease in the COVID-19 incidence rate. The accessibility of the test site had a negative impact on the COVID-19 incidence rate, and this impact was statistically significant in the results of OLS and SEM. For every one-point increase in test site accessibility, the COVID-19 incidence rate decreases by 75.207, as the results of SEM showed.

Rate of the population older than 65 was negatively associated with the COVID-19 incidence rate, as demonstrated by the result of OLS, but the results of two spatial regression models showed that this relationship was positive. As to the rate of households with more than 1 occupants per room, the results of the three models were all significantly positive. Specifically, the result of SEM indicated that a one-point increase in the rate of households with more than 1 occupants per room is associated with a 157.385-point increase in COVID-19 incidence rate. With a parameter estimate of -66.217 (SEM), the rate of the population below the poverty level had a significantly negative influence on the COVID-19 incidence rate. The association between this variable and the COVID-19 incidence rate was also negative in OLS and SLM, although the relationship had no statistical significance in the result of SLM. The results of the three models demonstrated that income inequality had a nonsignificant and negative impact on the COVID-19 incidence rate. Road density had a strong positive influence on the COVID-19 incidence rate, and the influence was statistically significant, as demonstrated by the results of three models. A one-point increase in road density is associated with a 226.89-point increase in COVID-19 incidence rate, according to the result of SEM.

## 5. Discussion

Racial/ethnic segregation is regarded as a fundamental cause of disparities in diseases (Williams and Collins, 2001). This study investigates the association between racial segregation and the COVID-19 incidence rate in Massachusetts, particularly the minority groups, such as Hispanic, Black/African American, and Asian. We find higher Hispanic and Black/African American segregations are more likely to be associated with a higher incidence rate. The areas where many black people reside are in poor areas characterized by high housing densities (Yancy, 2020). As revealed in the regression model, a higher percentage of more than 1 occupants per room and a higher poverty rate are significantly associated with the incidence rate as well. The higher observed incidence and severity in minority groups may be also associated with socioeconomic, cultural, or lifestyle factors, genetic predisposition, or pathophysiological differences in susceptibility or response to infection (Khunti, 2020). Bhala (2020) also proposed the important role of culture, including multigenerational households, and variation in social interactions have a role in the increased risk of COVID-19. Moreover, minorities also contribute to heightened exposure to COVID-19. They are more likely to work in industries that have remained open during non-essential business closures (Raifman, 2020).

The racial disparities in medical care treatments and outcomes are pervasive (Smedley et al., 2002). In Massachusetts, Black and Hispanic residents were less likely to be insured than white residents and they were more likely to report fair or poor health than whites (Zhu et al., 2010). Server disease increases the chance of being infected by COVID-19. For example, Yang et al. (2020) conducted a meta-analysis of eight studies including 46,248 patients with laboratory-confirmed cases of COVID-19. It indicated that those with the most severe disease were more likely to have hypertension, respiratory disease, and cardiovascular disease. Moreover, other studies found obesity and smoking were associated with increased risks (Huang, 2020; Wang 2020). In Italy, higher risks have also been reported in men than in women, which could be partly due to their higher smoking rates and subsequent comorbidities (Livingston, 2020). The pandemic presents a window of opportunity for achieving greater equity in the health care of all vulnerable populations (Hooper, 2020).

The access to testing sites shows Black population has the shortest drive time of 5.69 minutes to testing sites, followed by Hispanic, Asian, and Whites, which is consistent with those to hospitals in travel times of 9.22, 8.88, 9.96, and 11.21 minutes. All travel times to two kinds of sites for COVID-19 testing are acceptable. Such findings show some discrepancies addressed in previous studies. Yancy (2020) demonstrated that African Americans have higher rates of COVID-19 infection and death. This case is not found in Massachusetts as it has a lower Black population of 9% and 9.4% COVID-19 confirmed cases while the White is dominant with 80% population and 29.6 % confirmed cases. Another research stated COVID-19 testing centers are more likely to be established in the well-off suburbs of white dominates than in low-income black dominant neighborhoods (Williams and Copper, 2020). In fact, most testing sites and hospitals of Massachusetts are distributed in neighborhoods dominated by minorities as they tend to live for more job opportunities and accessible public transportation. The longer travel time of the White population to access testing sites and hospitals may be attributable to their preference of living far from cities for spacious houses. In addition, good access to testing sites might not be the only factor to influence the COVID-19 incidence rate but it is an important component to identify the possible carriers of COVID-19. Note that we assume car-driving is the only transport mode. The omission of other transport modes, such as public transit in well-developed regions might bias the estimation of travel times for testing.

Previous studies demonstrated that people over 65 are at the highest risk (Centers for Disease Control and Prevention, 2020), however, our study suggested that the relationship between COVID-19 incidence rate and the proportion of elderly people had no statistical significance. This may be attributable to the geographical distribution of the senior citizens in Massachusetts. As shown in Figure 4, cities/towns with high percentages of the elderly population are mainly located in the west and eastern coastal areas, which are far from the Greater Boston area.

**Figure 4.**
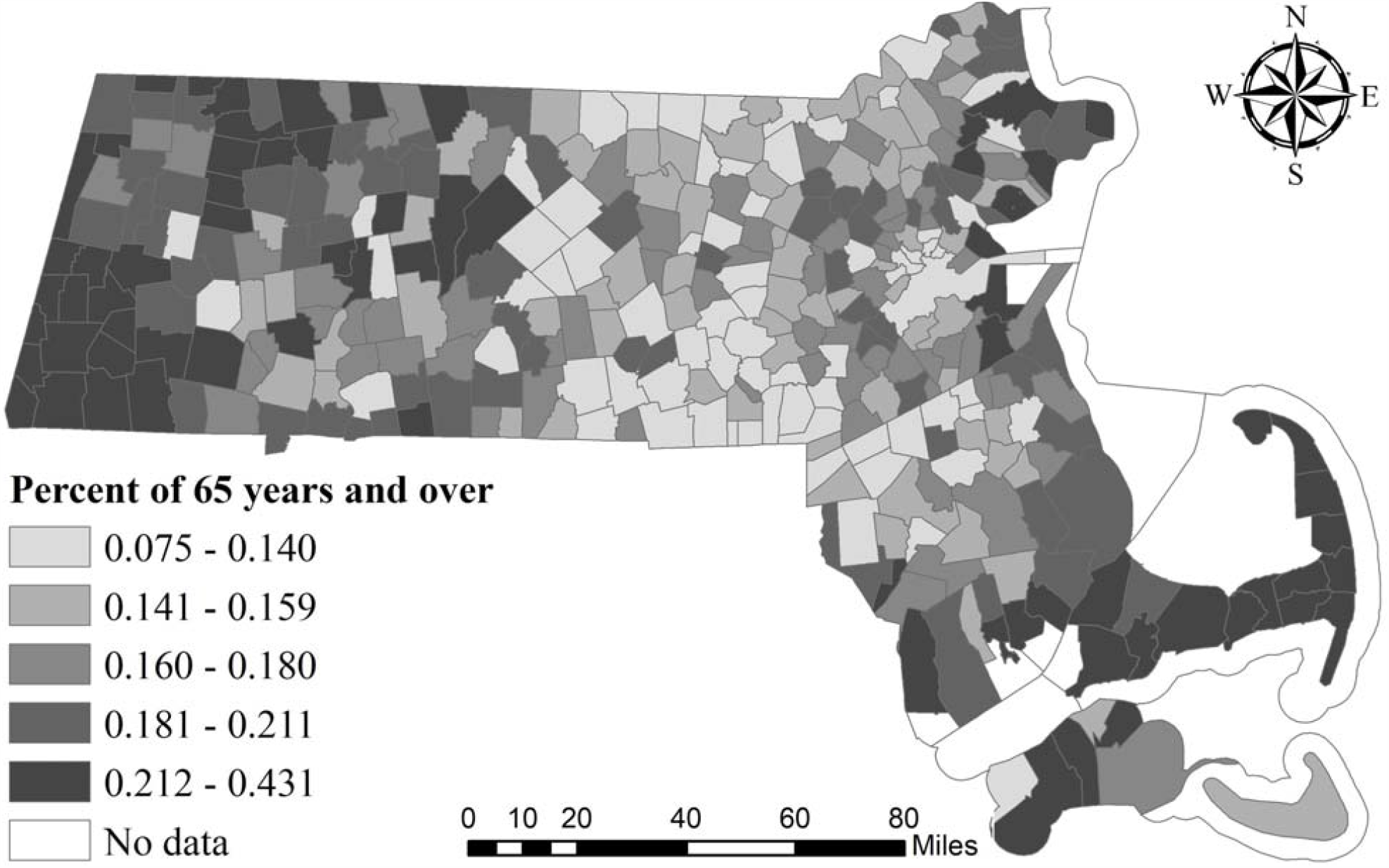
Geographical distribution of percentage of population 65 years and over.

Results of SEM suggested a strongly significant and positive relationship between COVID-19 incidence rate and percent of occupied housing units with more than 1 occupants per room. Quarantines are often one of the first responses against new infectious diseases. However, it is estimated that 44% of secondary cases were infected during the index cases’ presymptomatic stage, in settings with substantial household clustering, and quarantine outside the home (He, 2020). Several people sharing the same room in households with lower income may increase the chance for virus transmission. This study validated this hypothesis by showing that a higher percentage of more than 1 occupants per room and higher poverty rates are significantly associated with the incidence rate. The results of SEM also suggested a strongly significant and negative impact of the poverty rate on the COVID-19 incidence rate in Massachusetts. At first glance, this result is counterintuitive, since those in poverty are generally at risk of losing their health insurance coverage, which makes them vulnerable in the face of emergent epidemics like COVID-19. However, most cities/towns with high poverty rates are located in peripheral areas, which are remote from high COVID-19 incidence regions, as can be seen from Figure 5. Road density is significantly positively associated with COVID-19 incidence rate, this may be because cities/towns with higher road densities usually have more frequent human activities, which could provide conditions for disease transmission.

**Figure 5.**
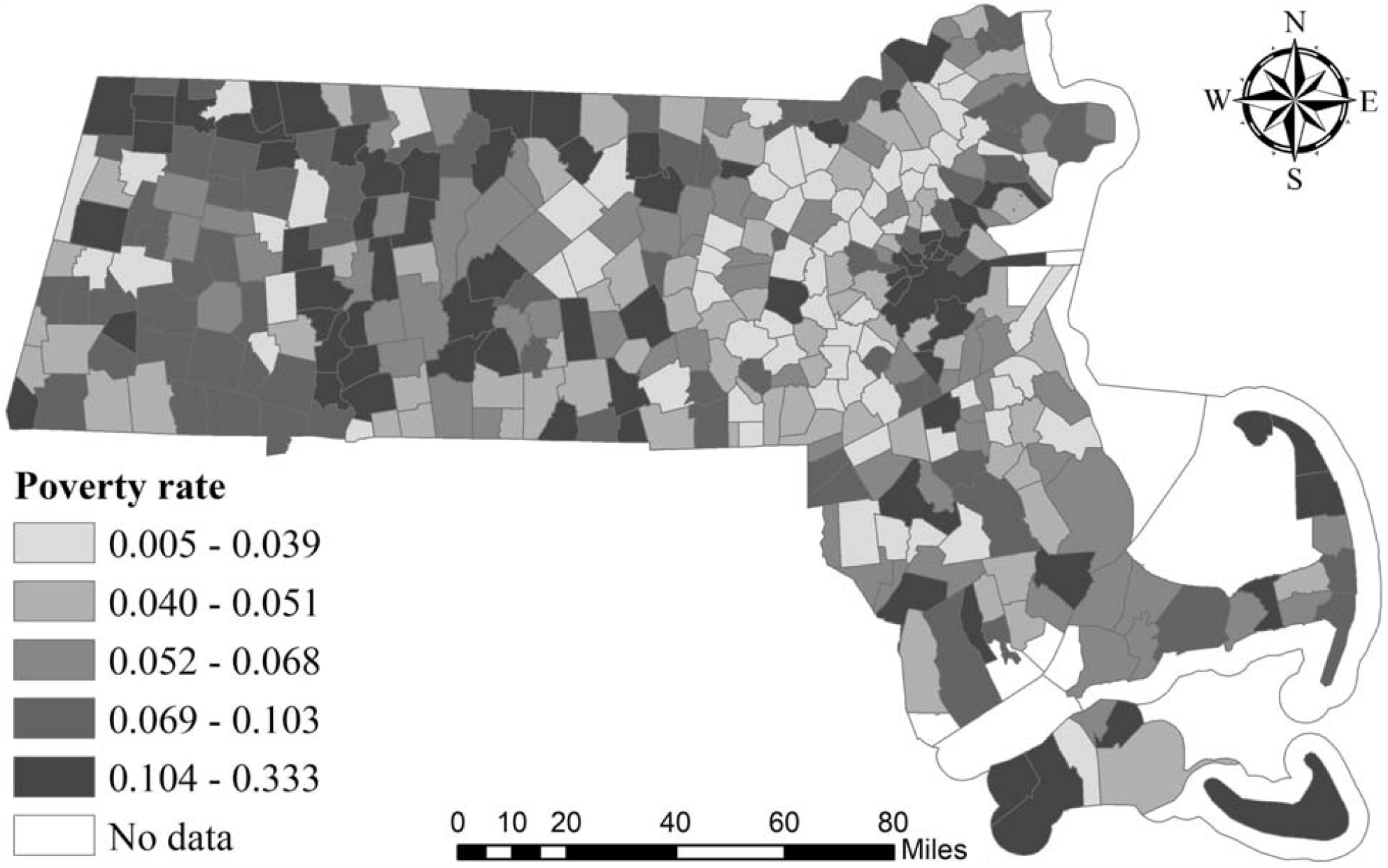
Geographical distribution of poverty rate.

## 6. Conclusions

The geographic disparity of the COVID-19 incidence rate has been recognized in previous studies (Hu et al., 2020; Liu et al., 2020; Yang et al., 2020). However, most research was conducted at or above the county level, which prevents us from observing how the COVID-19 incidence rate interacts with the various factors at a finer spatial scale. This study analyzed how the COVID-19 incidence rate was associated with racial and health accessibility disparities, after controlling for the possible influences of a set of demographics, economic, and transportation factors. The study was conducted at the city/town level in the State of Massachusetts, which is one of the states hit hardest by the pandemic.

The classic OLS regression model is unable to deal with the spatial autocorrelation effect. Therefore, we conducted a further spatial regression analysis based on two models, i.e., SLM and SEM, to obtain more robust estimation of the significances and directions of the influences of racial and health accessibility disparities, demographic, economic and transportation characteristics on COVID-19 incidence rate. Results suggest that residential segregations of Hispanic and Non-Hispanic Black/African Americans are associated with an increased risk of COVID-19 infection. Similarly, road density and the percent of more than 1 occupants per room have statistically significant and positive impacts on the COVID-19 incidence rate. However, test site accessibility and poverty rate are related to a decreased risk of COVID-19 infection.

The empirical findings shown in this paper could provide helpful insight and guidance for policymakers to develop strategies to contain the COVID-19 transmissions in Massachusetts. Firstly, political action is needed to resolve long-standing societal inequalities, addressing the injustices of public health, and tackling the COVID-19 pandemic and its sequence (Bhala, 2020). Public health is complicated and social reengineering is complex, but a change of this magnitude does not happen without a new resolve (Yancy, 2020). Additionally, as road density and the overcrowding are found to have so much influence on the pandemic transmission, then the social distancing or stay-at-home policies should be rigorously followed. As the condition of COVID-19 is still going on and evolving quickly in the United States, it is important to explore the spatial patterns in small units, such as neighborhoods. The research framework and implications of this study can also provide a reference for other areas to help understand the local transmission of COVID-19 and minimize the damage of the terrible disease.

## Data Availability

All data referred to in the manuscript is accessible with the website links provided in our references section.

